# REACT-1 round 8 interim report: SARS-CoV-2 prevalence during the initial stages of the third national lockdown in England

**DOI:** 10.1101/2021.01.20.21250158

**Authors:** Steven Riley, Haowei Wang, Oliver Eales, Caroline E. Walters, Kylie E. C. Ainslie, Christina Atchison, Claudio Fronterre, Peter J. Diggle, Deborah Ashby, Christl A. Donnelly, Graham Cooke, Wendy Barclay, Helen Ward, Ara Darzi, Paul Elliott

**Affiliations:** School of Public Health, Imperial College London, UK; MRC Centre for Global infectious Disease Analysis and Abdul Latif Jameel Institute for Disease and Emergency Analytics, Imperial College London, UK; CHICAS, Lancaster Medical School, Lancaster University, UK and Health Data Research, UK; Department of Statistics, University of Oxford, UK; Department of Infectious Disease, Imperial College London, UK; Imperial College Healthcare NHS Trust, UK; National Institute for Health Research Imperial Biomedical Research Centre, UK; Institute of Global Health Innovation at Imperial College London, UK; MRC Centre for Environment and Health, School of Public Health, Imperial College London, UK; Health Data Research (HDR) UK London at Imperial College; UK Dementia Research Institute at Imperial College; Centre for Infectious Disease Control, National Institute for Public Health and the Environment, Bilthoven, The Netherlands

**Author notes:** Corresponding authors: Steven Riley and Paul Elliott, School of Public Health, Imperial College London, Norfolk Place, London, W2 1PG.

## Abstract

**Background:** High prevalence of SARS-CoV-2 virus in many northern hemisphere populations is causing extreme pressure on healthcare services and leading to high numbers of fatalities. Even though safe and effective vaccines are being deployed in many populations, the majority of those most at-risk of severe COVID-19 will not be protected until late spring, even in countries already at a more advanced stage of vaccine deployment.

**Methods:** The REal-time Assessment of Community Transmission study-1 (REACT-1) obtains throat and nose swabs from between 120,000 and 180,000 people in the community in England at approximately monthly intervals. Round 8a of REACT-1 mainly covers a period from 6th January 2021 to 15th January 2021. Swabs are tested for SARS-CoV-2 virus and patterns of swab-positivity are described over time, space and with respect to individual characteristics. We compare swab-positivity prevalence from REACT-1 with mobility data based on the GPS locations of individuals using the Facebook mobile phone app. We also compare results from round 8a with those from round 7 in which swabs were collected from 13th November to 24th November (round 7a) and 25th November to 3rd December 2020 (round 7b).

**Results:** In round 8a, we found 1,962 positives from 142,909 swabs giving a weighted prevalence of 1.58% (95% CI, 1.49%, 1.68%). Using a constant growth model, we found no strong evidence for either growth or decay averaged across the period; rather, based on data from a limited number of days, prevalence may have started to rise at the end of round 8a. Facebook mobility data showed a marked decrease in activity at the end of December 2020, followed by a rise at the start of the working year in January 2021. Between round 7b and round 8a, prevalence increased in all adult age groups, more than doubling to 0.94% (0.83%, 1.07%) in those aged 65 and over. Large household size, living in a deprived neighbourhood, and Black and Asian ethnicity were all associated with increased prevalence. Both healthcare and care home workers, and other key workers, had increased odds of swab-positivity compared to other workers.

**Conclusion:** During the initial 10 days of the third COVID-19 lockdown in England in January 2021, prevalence of SARS-CoV-2 was very high with no evidence of decline. Until prevalence in the community is reduced substantially, health services will remain under extreme pressure and the cumulative number of lives lost during this pandemic will continue to increase rapidly.

## Introduction

As the COVID-19 pandemic enters 2021, northern hemisphere populations that have not achieved ongoing containment [1] face substantial challenges. Although effective vaccines are available [2,3], it will be many weeks until a substantial proportion of those most at-risk of severe disease are protected, even in countries such as England where large-scale vaccine roll-out for the most vulnerable groups has been implemented [4]. Prevalence of infection at the beginning of 2021 is at the highest levels [5] since the peak of the first wave in March and April 2020 [6]. There has been a sharp increase in infections since the beginning of the second wave in autumn 2020, and, notwithstanding some fluctuations in prevalence coinciding e.g. with school half-term holidays and the second lockdown in England, prevalence has been high for many weeks [7]. This has inevitably led to extreme demand for health services and daily numbers of deaths as high or higher than the first wave of the pandemic [8]. Furthermore, SARS-CoV-2 is evolving epidemiologically into distinct lineages, with potentially increased transmissibility [9] and significant antigenic changes [10].

England responded to the current situation with its third national lockdown, starting on 6th January 2021 [11]. People are asked to: stay at home whenever possible (including working from home), only to do essential shopping and to meet outside only with one other person from other households.

Here, we present results from the latest round (8a) of the REACT-1 study which commenced self-administered swab-collection from large numbers of people on 6th January 2021 (though a small number of tests were obtained from the 30th December). We report results from swabs collected up to and including 15th January (with a small number from subsequent days) and compare the results with those from round 7 in which swabs were collected from 13th November to 24th November (round 7a) and 25th November to 3rd December 2020 (round 7b).

## Results

In round 8a, we found 1,962 positives from 142,909 swabs giving a weighted prevalence of 1.58% (95% confidence interval, 1.49%, 1.68%) (Table 1). This is the highest prevalence recorded by REACT-1 since it started in May 2020 and represents a greater than 50% increase from 0.91% (0.81%, 1.03%) observed during round 7b (when prevalence was increasing). Since there was a gap of over one month from the end of round 7b on 3rd December 2020 to beginning of round 8a on 6th January 2021 we may have missed a peak in prevalence during the intervening period.

**Table 1.** Unweighted and weighted prevalence of swab-positivity across seven rounds of REACT-1.

| Round | Tested swabs | Positive swabs | Unweighted prevalence (95% CI) | Weighted prevalence (95% CI) | First sample | Last sample |
| --- | --- | --- | --- | --- | --- | --- |
| 1 | 120,620 | 159 | 0.13% (0.11%, 0.15%) | 0.16% (0.13%, 0.19%) | 1/5/2020 | 1/6/2020 |
| 2 | 159,199 | 123 | 0.077% (0.065%, 0.092%) | 0.088% (0.068%, 0.11%) | 19/6/2020 | 7/7/2020 |
| 3 | 162,821 | 54 | 0.033% (0.025%, 0.043%) | 0.040% (0.027%, 0.053%) | 24/7/2020 | 11/8/2020 |
| 4 | 154,325 | 137 | 0.089% (0.075%, 0.11%) | 0.13% (0.096%, 0.15%) | 20/8/2020 | 8/9/2020 |
| 5 | 174,949 | 824 | 0.47% (0.44%, 0.50%) | 0.60% (0.55%, 0.71%) | 18/9/2020 | 5/10/2020 |
| 6 | 160,175 | 1,732 | 1.08% (1.03%, 1.13%) | 1.30% (1.21%, 1.39%) | 16/10/2020 | 2/11/2020 |
| 6a | 85,965 | 863 | 1.00% (0.94%, 1.07%) | 1.28% (1.16%, 1.42%) | 16/10/2020 | 25/10/2020 |
| 6b | 74,210 | 869 | 1.17% (1.10%, 1.25%) | 1.32% (1.20%, 1.45%) | 26/10/2020* | 2/11/2020 |
| 7 | 168,181 | 1,299 | 0.77% (0.73%, 0.82%) | 0.94% (0.87%, 1.01%) | 13/11/2020 | 3/12/2020 |
| 7a | 105,122 | 821 | 0.78% (0.73%, 0.84%) | 0.96% (0.87%, 1.05%) | 13/11/2020 | 24/11/2020 |
| 7b | 63,059 | 478 | 0.76% (0.69%, 0.83%) | 0.91% (0.81%, 1.03%) | 25/11/2020* | 3/12/2020 |
| 8a | 142,909 | 1,962 | 1.37% (1.31%, 1.43%) | 1.58% (1.49%, 1.68%) | 06/01/2021** | 15/01/2021*** |

\* Includes some samples from prior period
\*\* Includes some swab tests from 30th December onwards
\*\*\* Includes small number of samples from subsequent days

Using a constant growth model, we found no strong evidence for either growth or decay averaged across the period of round 8a (Table 2, Figure 1), with estimated reproduction number (R) at 1.04 (0.94, 1.15). However, we found evidence for non-constant growth within round 8a; a logistic regression model with a smooth term gave ΔAIC > 6 compared with one using only a constant term. Fitting a p-spline to the data across all rounds suggested prevalence may have been declining at start of round 8a, from an unobserved earlier peak. Based on data from a limited number of days, prevalence may have started to rise at the end of round 8a (Figure 2). Using regional p-splines, sub-national prevalence was consistent with the national pattern of a recent plateau or increase, other than in the South West, where prevalence was decreasing (Table 3, Figure 3).

**Table 2.** Estimates of national growth rates, doubling times and reproduction numbers for round 8a.

| Round | Outcome | Growth rate | R* | Probability R>1 |
| --- | --- | --- | --- | --- |
| 8a | All positives | 0.007 ( -0.010 , 0.023 ) | 1.04 ( 0.94 , 1.15 ) | 0.79 |
|  | Non-symptomatics | -0.007 ( -0.037 , 0.022 ) | 0.96 ( 0.78 , 1.14 ) | 0.33 |
|  | Positive for both E and N genes | -0.004 ( -0.024 , 0.017 ) | 0.98 ( 0.85 , 1.11 ) | 0.36 |
|  | Positive for both E and N genes or positive only for N gene with CT 35 or less | 0.000 ( -0.018 , 0.018 ) | 1.00 ( 0.89 , 1.12 ) | 0.52 |

\* Doubling/Halving time not shown because R close to 1

**Table 3.**
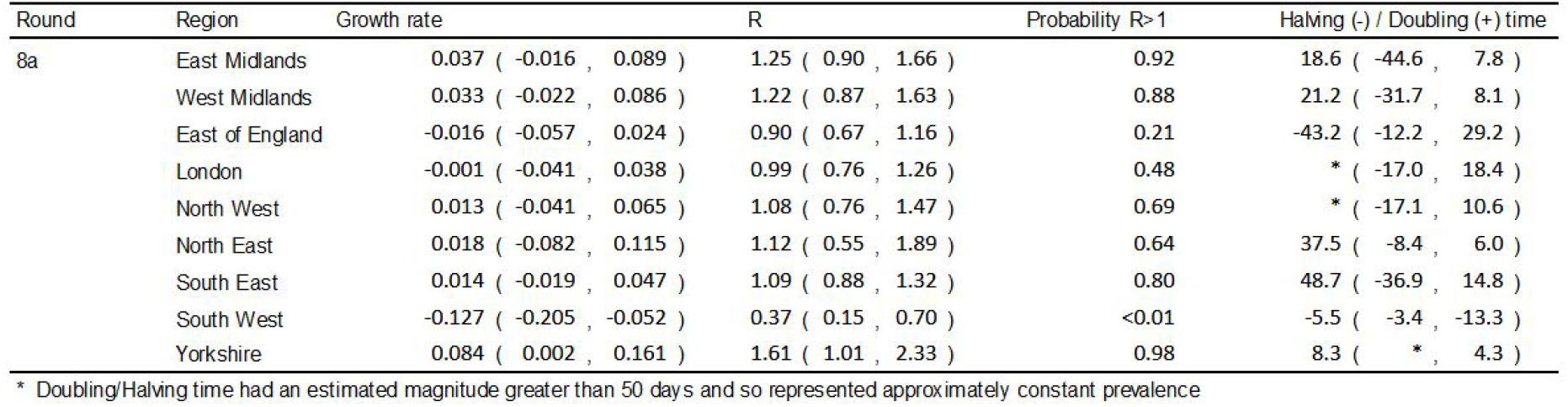
Estimates of regional growth rates, doubling times and reproduction numbers for round 8a.

| Round | Region | Growth rate | R | Probability R>1 | Halving (-) / Doubling (+) time |
| --- | --- | --- | --- | --- | --- |
| 8a | East Midlands | 0.037 ( -0.016 , 0.089 ) | 1.25 ( 0.90 , 1.66 ) | 0.92 | 18.6 ( -44.6 , 7.8 ) |
|  | West Midlands | 0.033 ( -0.022 , 0.086 ) | 1.22 ( 0.87 , 1.63 ) | 0.88 | 21.2 ( -31.7 , 8.1 ) |
|  | East of England | -0.016 ( -0.057 , 0.024 ) | 0.90 ( 0.67 , 1.16 ) | 0.21 | -43.2 ( -12.2 , 29.2 ) |
|  | London | -0.001 ( -0.041 , 0.038 ) | 0.99 ( 0.76 , 1.26 ) | 0.48 | * ( -17.0 , 18.4 ) |
|  | North West | 0.013 ( -0.041 , 0.065 ) | 1.08 ( 0.76 , 1.47 ) | 0.69 | * ( -17.1 , 10.6 ) |
|  | North East | 0.018 ( -0.082 , 0.115 ) | 1.12 ( 0.55 , 1.89 ) | 0.64 | 37.5 ( -8.4 , 6.0 ) |
|  | South East | 0.014 ( -0.019 , 0.047 ) | 1.09 ( 0.88 , 1.32 ) | 0.80 | 48.7 ( -36.9 , 14.8 ) |
|  | South West | -0.127 ( -0.205 , -0.052 ) | 0.37 ( 0.15 , 0.70 ) | <0.01 | -5.5 ( -3.4 , -13.3 ) |
|  | Yorkshire | 0.084 ( 0.002 , 0.161 ) | 1.61 ( 1.01 , 2.33 ) | 0.98 | 8.3 ( * , 4.3 ) |

\* Doubling/Halving time had an estimated magnitude greater than 50 days and so represented approximately constant prevalence

**Figure 1.**
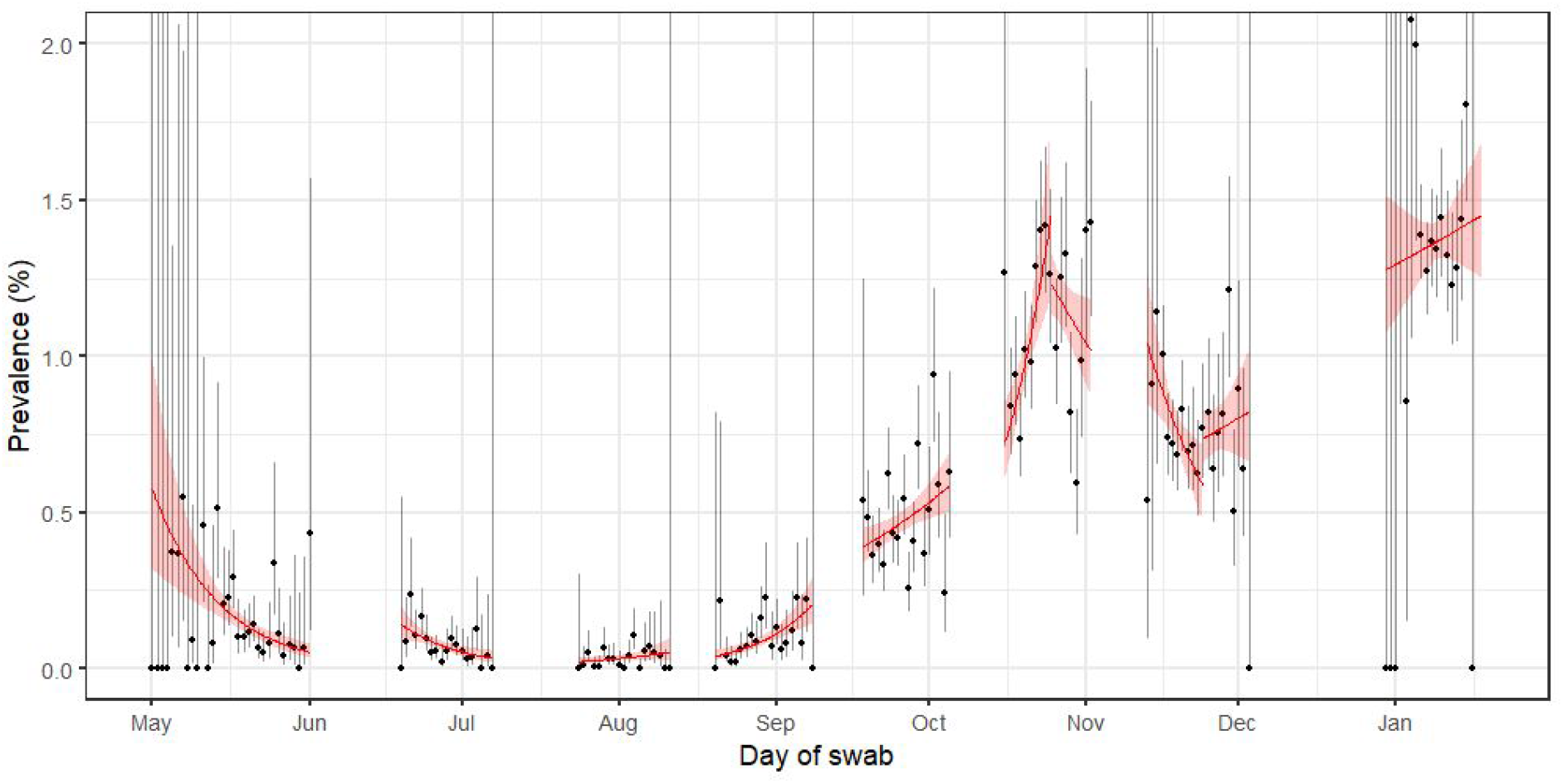
Constant growth rate models fit to REACT-1 data for England for individual rounds (and partial rounds for rounds 6 and 7). Shaded area shows the central 95% posterior credible interval.

**Figure 2.**
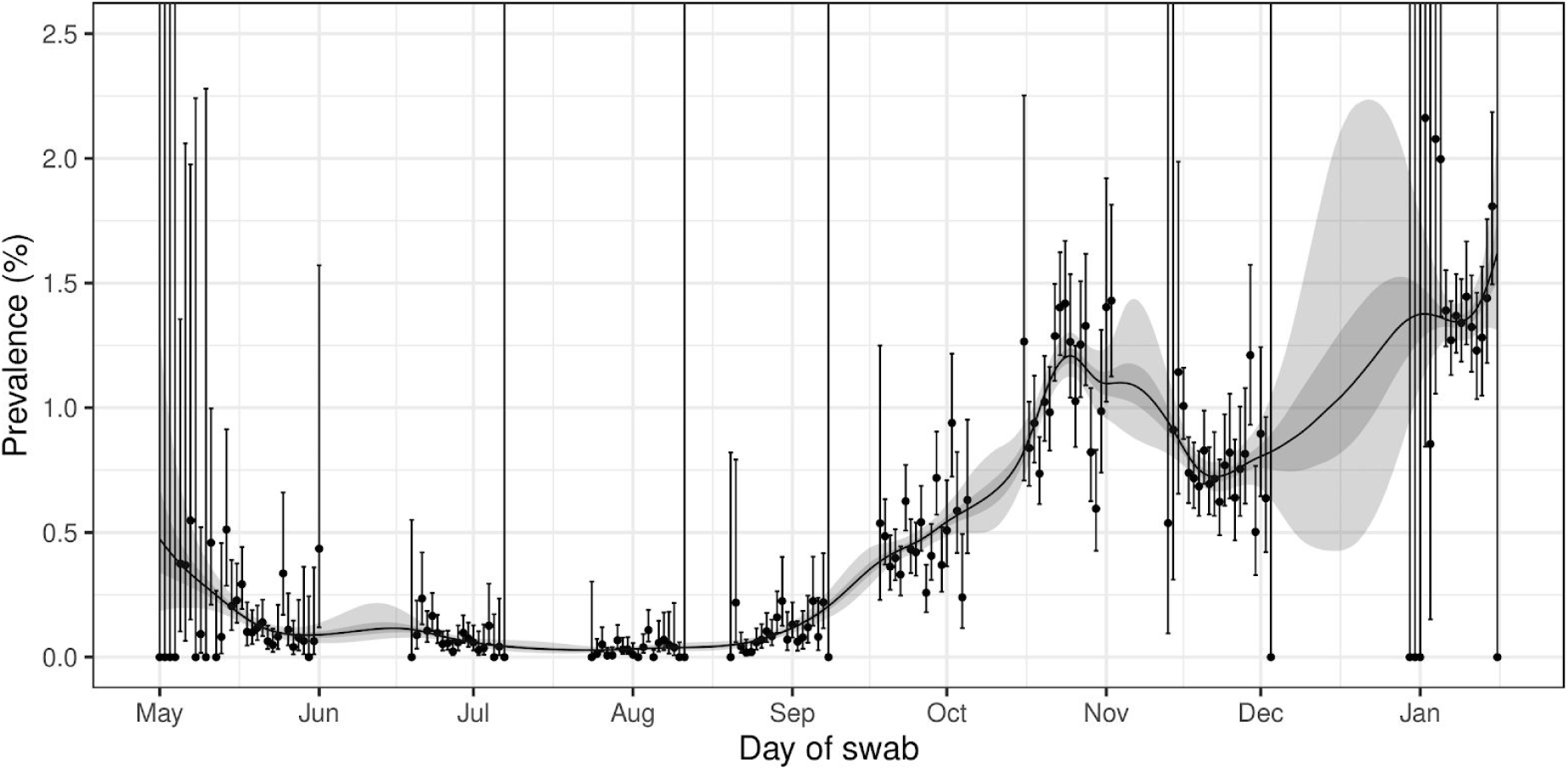
Prevalence of national swab-positivity for England estimated using a p-spline for the full period of the study with central 50% (dark grey) and 95% (light grey) posterior credible intervals.

**Figure 3.**
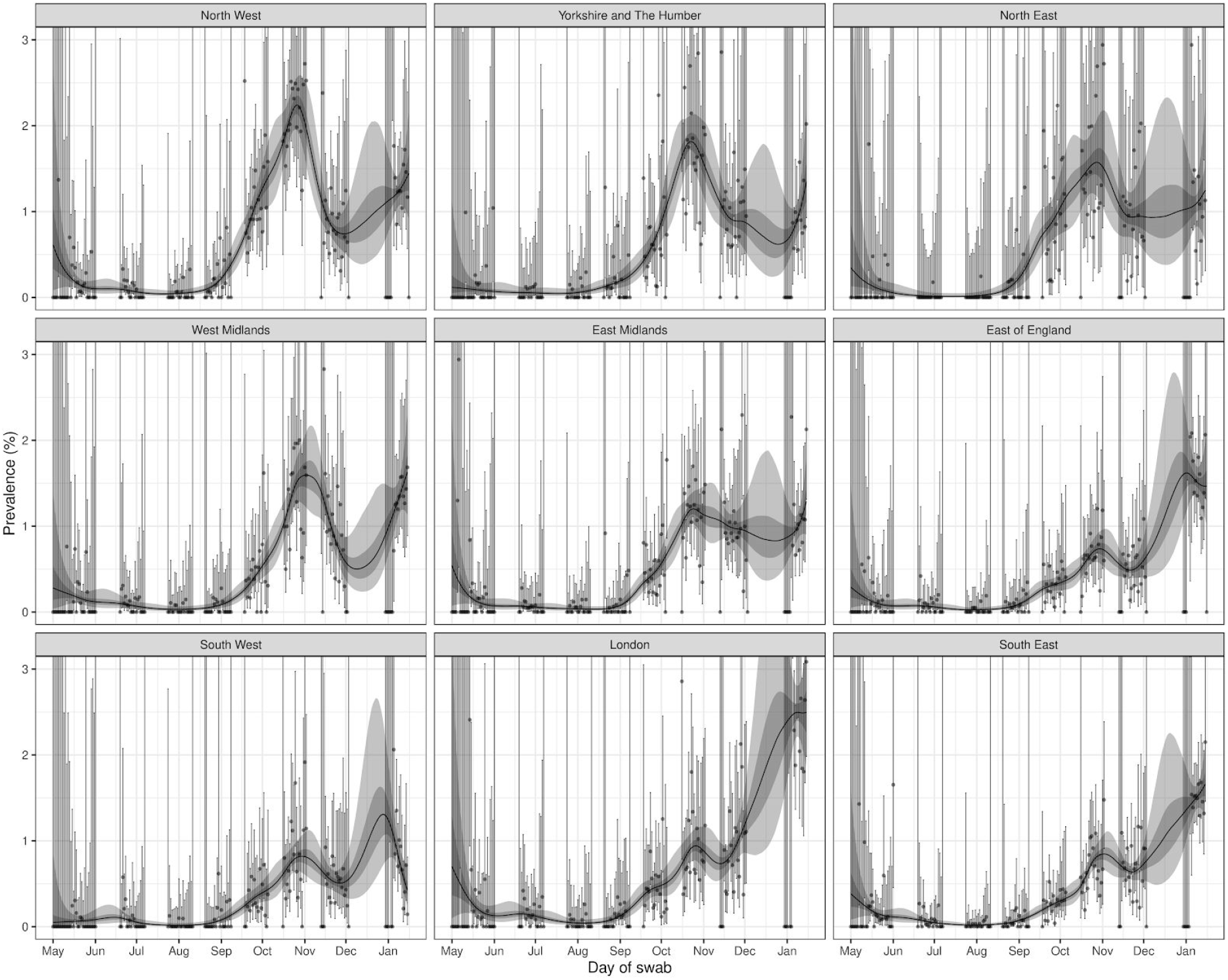
Prevalence of swab-positivity for each region estimated using a p-spline (with a constant second-order random walk prior distribution) for the full period of the study with central 50% (dark grey) and 95% (light grey) posterior credible intervals.

Mobility data from the Facebook app suggested that there was a marked decrease in activity nationally at the end of December 2020, followed by a rise at the start of the working year in January 2021 (Figure 4). These mobility patterns may help explain changes in prevalence observed during January 2021.

**Figure 4.**
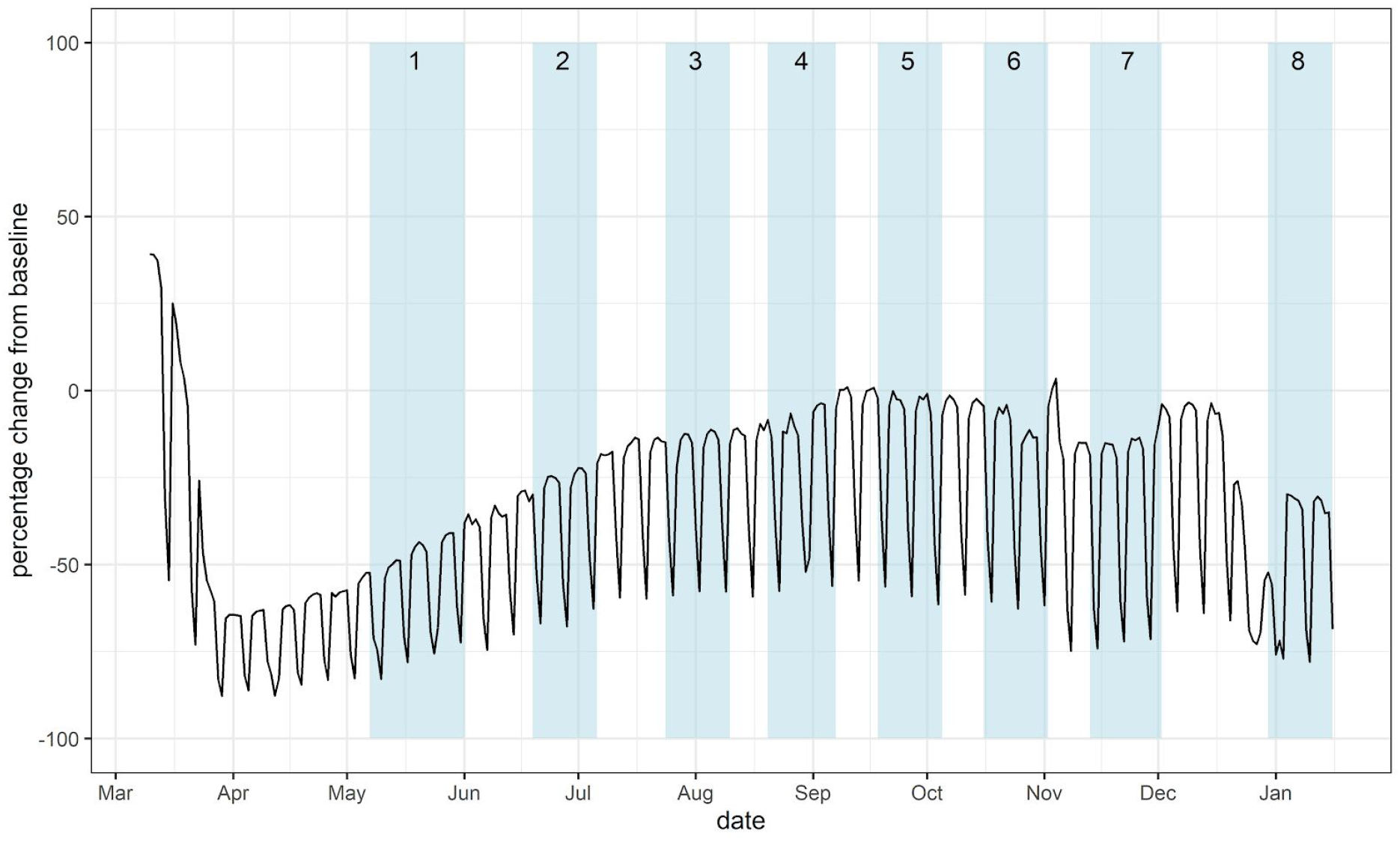
Facebook mobility data for England (black line) and the REACT-1 rounds (blue shaded regions). Baseline for mobility data is the mean movement during the week 10th - 16th March 2020 inclusive.

We observed changes in weighted prevalence at the regional scale (Table 4, Figure 5) between rounds 7b and 8a. Prevalence in round 8a was highest in London at 2.80% (2.47%, 3.17%), more than a doubling from 1.21% (0.91%, 1.59%) in round 7b. Prevalence also more than doubled in: South East from 0.75% (0.59%, 0.95%) to 1.68% (1.51%, 1.87%); East of England from 0.59% (0.43%, 0.81%) to 1.74% (1.52%, 1.99%); and West Midlands from 0.71% (0.43%, 1.16%) to 1.76% (1.37%, 2.26%). In addition, prevalence increased in the South West from 0.53% (0.36%, 0.78%) to 0.94% (0.75%, 1.19%) and there was an apparent increase in North West. For the other regions, there was an apparent decrease in prevalence in Yorkshire and The Humber while prevalence was broadly similar (comparing rounds 7b and 8a) in East Midlands and North East.

**Table 4.**
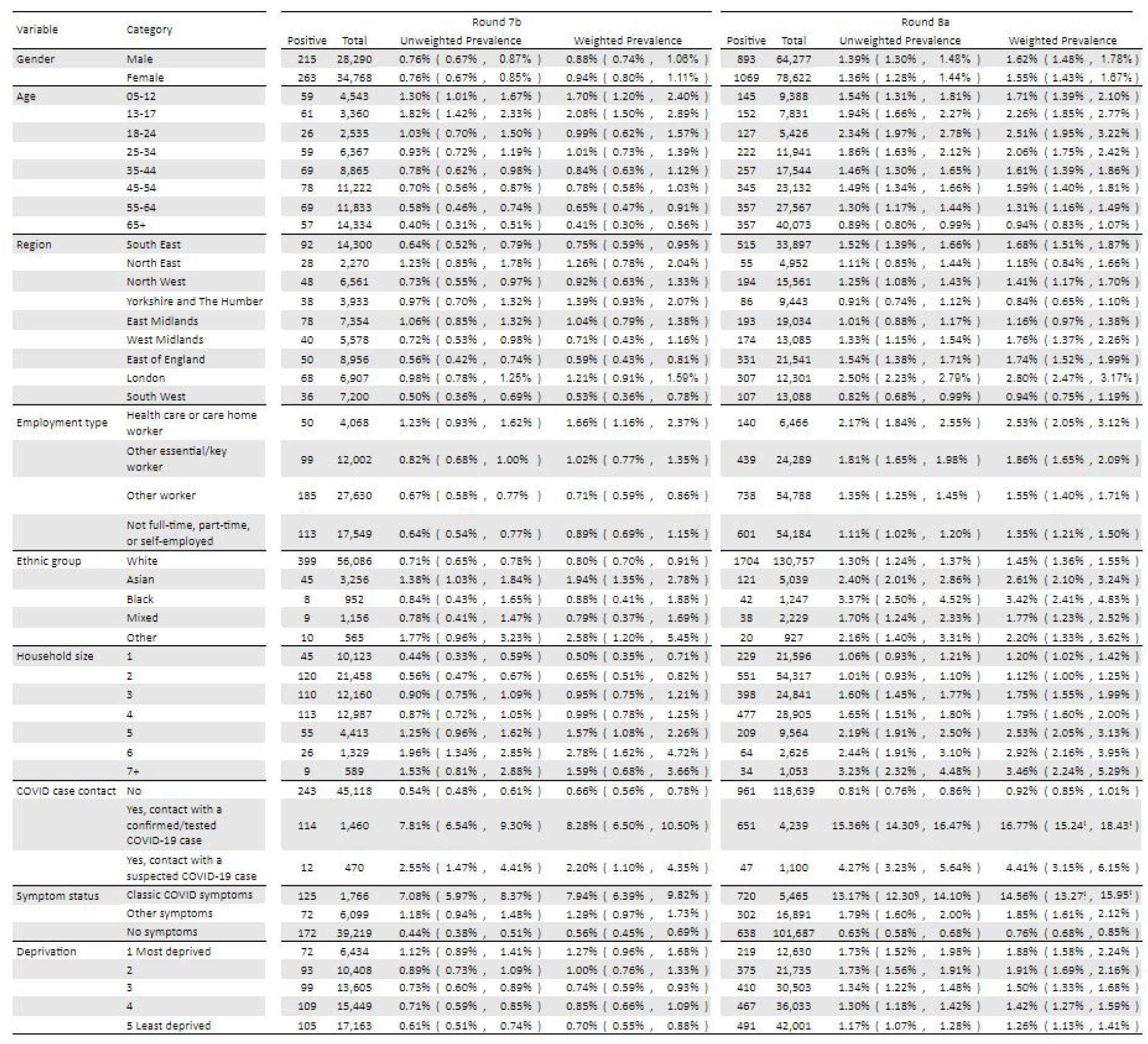
Unweighted and weighted prevalence of swab-positivity for rounds 7b and 8a.

**Figure 5.**
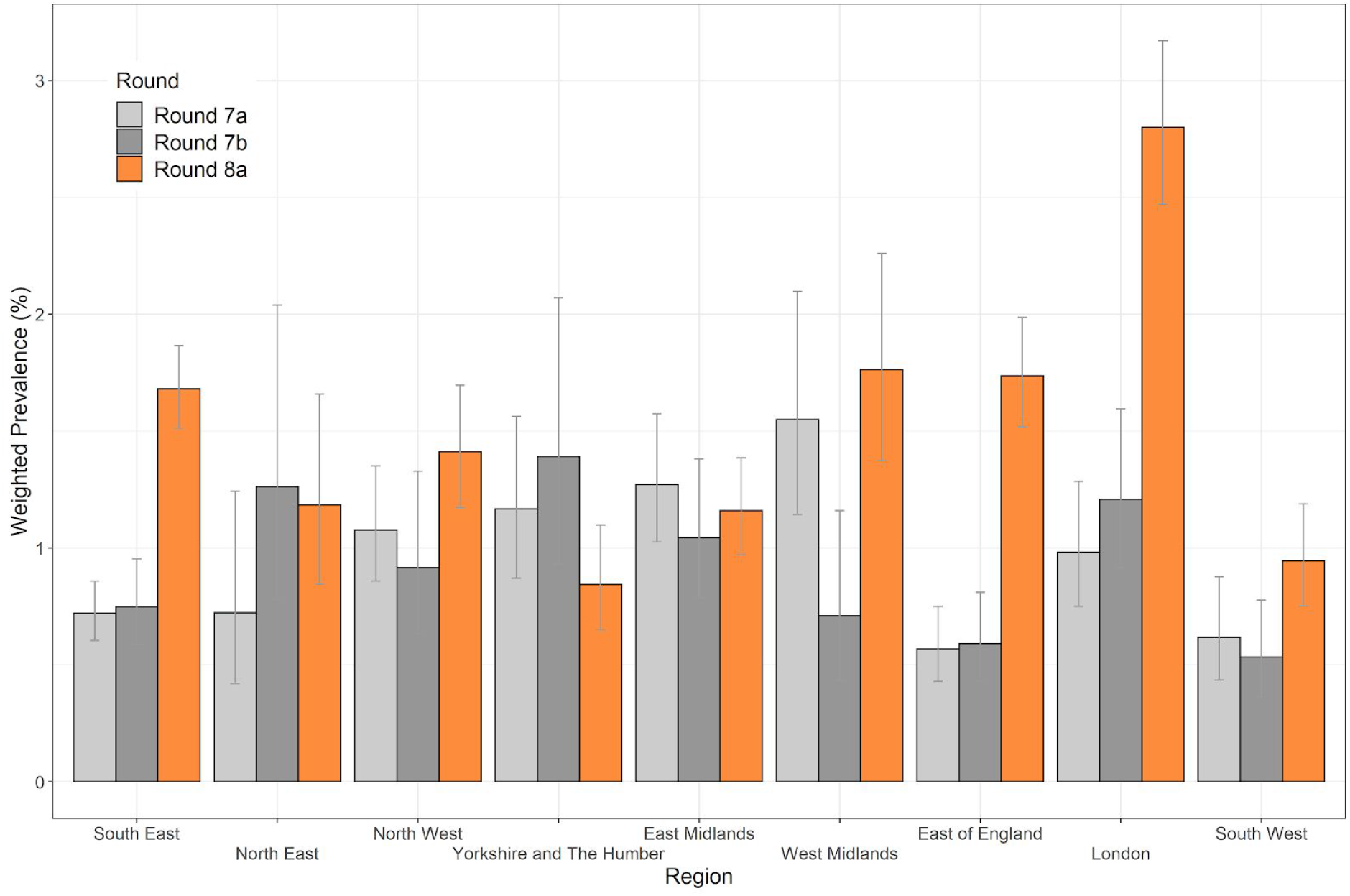
Weighted prevalence of swab-positivity by region for rounds 7a, 7b and 8a. Bars show 95% confidence intervals.

We also investigated patterns by age and showed that between rounds 7b and 8a, weighted prevalence increased in all adult age groups (Table 4, Figure 6). It was highest in 18 to 24 year olds with a weighted prevalence of 2.51% (1.95%, 3.22%), while prevalence in those aged 65 and over more than doubled to 0.94% (0.83%, 1.07%) in round 8a from 0.41% (0.30%, 0.56%) in round 7b.

**Figure 6.**
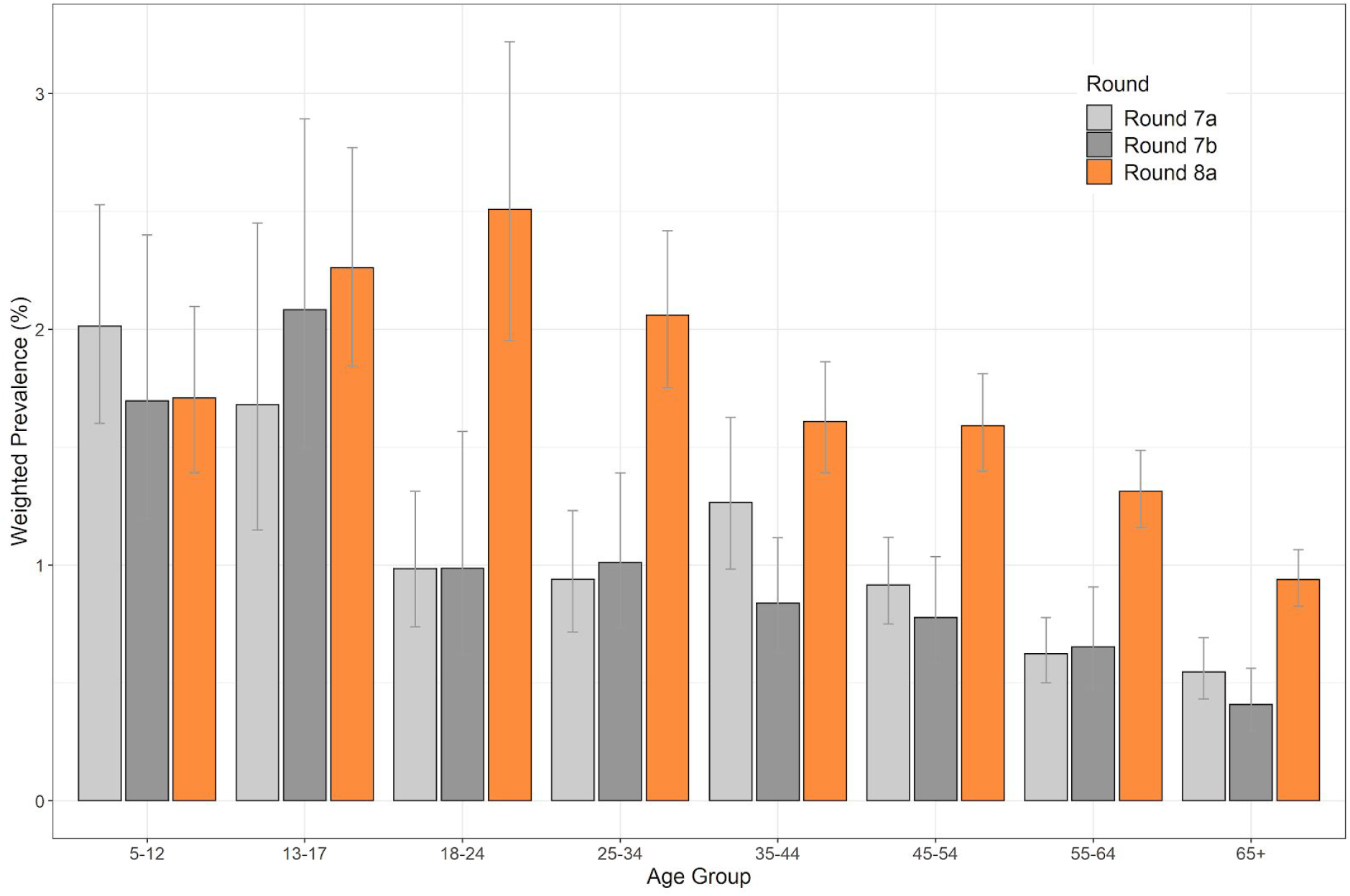
Weighted prevalence of swab-positivity by age group for rounds 7a, 7b and 8a. Bars show 95% confidence intervals.

Despite some uncertainty, age-prevalence patterns varied substantially by region (Figure 7). From rounds 7b to 8a, we observed large increases at older ages in London, South East, and East of England. In round 8a, London had the highest weighted prevalence nationally at greater than 2% in those aged 55 to 64 and in those 65 years and over. In contrast, patterns in Yorkshire and The Humber, North East and East Midlands did not show increases between rounds 7b and 8a in older adult ages. In addition, we observed weighted prevalence of over 4% in London among those aged 18-24 years in round 8a.

**Figure 7.**
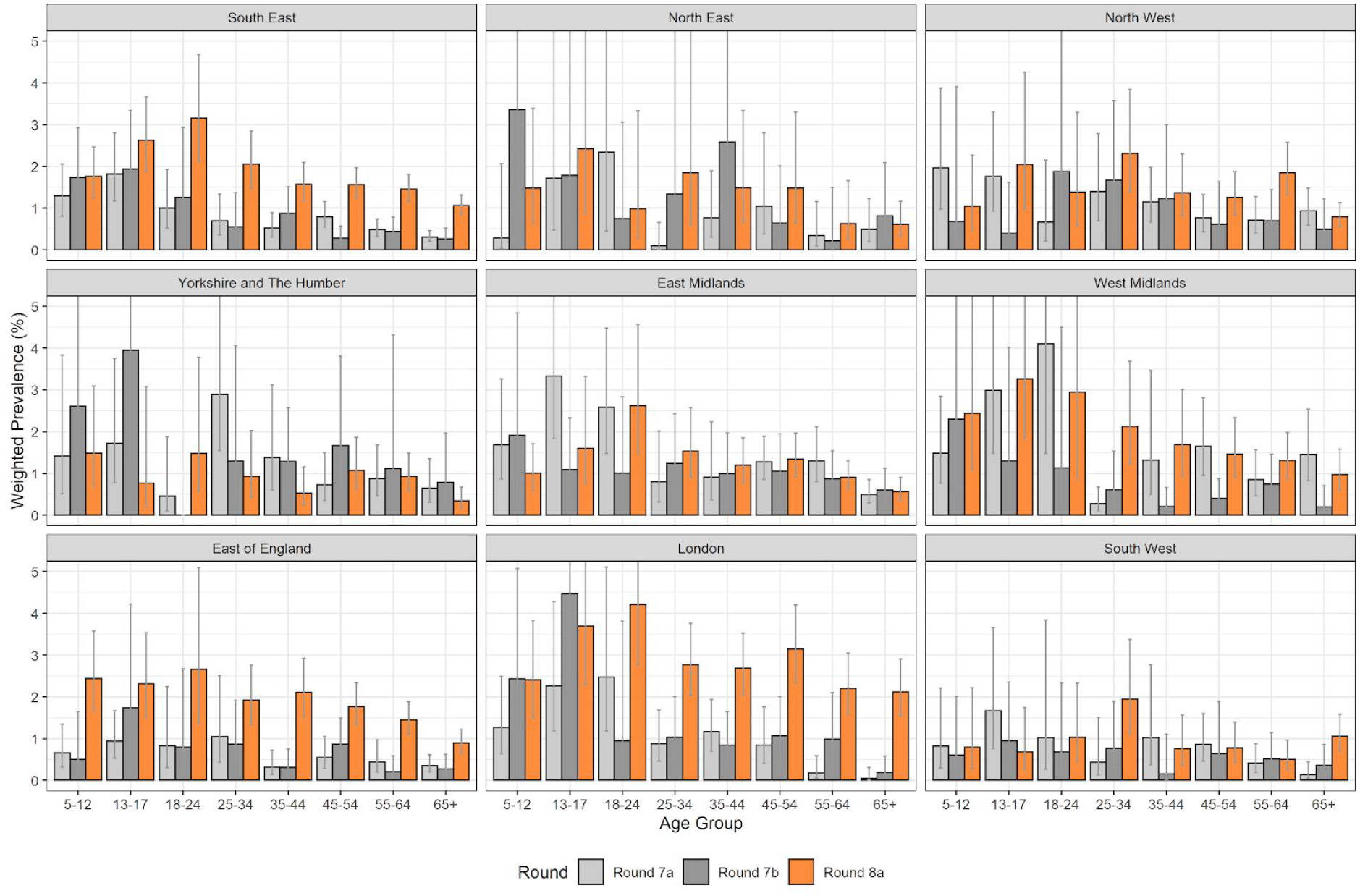
Weighted prevalence of swab-positivity by age group and region for rounds 7a, 7b, and 8a. Bars show 95% confidence intervals.

We found that large household size, living in a deprived neighbourhood, and Black and Asian ethnicity were all associated with increased prevalence (Table 4). We observed a monotonic increase in prevalence from the smallest to the largest households rising from 1.20% (1.02%, 1.42%) in single-person households to 3.46% (2.24%, 5.29%) in households of seven or more people. People living in neighbourhoods in the two most deprived quintiles had prevalence of 1.88% (1.58%, 2.24%) and 1.92% (1.69%, 2.16%) compared with 1.26% (1.13%, 1.41%) for those in the least deprived neighbourhoods. We found increased prevalence among participants of Black and Asian ethnicity at 3.42% (2.41%, 4.83%) and 2.61% (2.10%, 3.24%) respectively compared with 1.45% (1.36%, 1.55%) among white participants. Patterns of prevalence for household size, neighbourhood deprivation and ethnicity were reflected in elevated odds ratios for these variables in a jointly adjusted logistic regression model (Table 5, Figure 8).

**Table 5.** Jointly adjusted odds ratios for swab-positivity by: gender, age, region, key worker status, ethnicity, household size and index of area deprivation.

| Variable | Category | Round |  |  |
| --- | --- | --- | --- | --- |
|  |  | 7a | 7b | 8a |
| Gender | Male | Ref | Ref | Ref |
|  | Female | 0.92 [0.80,1.06] | 0.96 [0.79,1.16] | 0.94 [0.86,1.04] |
| Age Group | 5-12 | 1.46 [1.07,1.97] | 1.59 [1.10,2.29] | 0.99 [0.80,1.22] |
|  | 13-17 | 2.52 [1.80,3.53] | 2.27 [1.45,3.55] | 1.65 [1.30,2.09] |
|  | 18-24 | 2.05 [1.46,2.87] | 1.31 [0.82,2.11] | 1.72 [1.38,2.14] |
|  | 25-34 | 1.11 [0.80,1.52] | 1.30 [0.91,1.86] | 1.37 [1.14,1.65] |
|  | 35-44 | Ref | Ref | Ref |
|  | 45-54 | 1.18 [0.90,1.54] | 0.98 [0.70,1.37] | 1.14 [0.97,1.35] |
|  | 55-64 | 0.98 [0.73,1.31] | 0.95 [0.67,1.36] | 1.19 [1.00,1.42] |
|  | 65+ | 0.85 [0.61,1.17] | 0.74 [0.49,1.12] | 1.04 [0.85,1.26] |
| Region | North East | 0.99 [0.63,1.54] | 1.88 [1.20,2.95] | 0.69 [0.52,0.92] |
|  | North West | 1.30 [1.01,1.69] | 1.09 [0.75,1.58] | 0.77 [0.65,0.92] |
|  | Yorkshire | 1.42 [1.06,1.90] | 1.51 [1.01,2.25] | 0.60 [0.48,0.76] |
|  | East Midlands | 1.39 [1.10,1.77] | 1.63 [1.18,2.24] | 0.66 [0.55,0.78] |
|  | West Midlands | 1.64 [1.28,2.11] | 1.11 [0.75,1.64] | 0.85 [0.71,1.02] |
|  | East of England | 0.68 [0.51,0.90] | 0.92 [0.65,1.32] | 0.97 [0.84,1.12] |
|  | London | 0.89 [0.65,1.21] | 1.27 [0.90,1.80] | 1.44 [1.23,1.68] |
|  | South East | Ref | Ref | Ref |
|  | South West | 0.83 [0.60,1.15] | 0.81 [0.54,1.22] | 0.54 [0.43,0.66] |
| Key Worker Status | HCW/CHW | 1.69 [1.29,2.21] | 1.68 [1.22,2.32] | 1.66 [1.38,2.00] |
|  | Key worker (other) | 1.19 [0.98,1.45] | 1.14 [0.89,1.47] | 1.35 [1.20,1.53] |
|  | Other worker | Ref | Ref | Ref |
|  | Not FT, PT, SE | 0.85 [0.70,1.04] | 0.99 [0.76,1.30] | 0.90 [0.80,1.03] |
| Ethnicity | Asian | 1.70 [1.25,2.32] | 1.42 [1.01,2.01] | 1.26 [1.04,1.54] |
|  | Black | 1.23 [0.63,2.41] | 0.87 [0.42,1.78] | 1.61 [1.17,2.23] |
|  | Mixed | 0.98 [0.57,1.68] | 0.76 [0.38,1.55] | 1.02 [0.73,1.42] |
|  | Other | 1.40 [0.62,3.17] | 1.94 [0.98,3.82] | 1.20 [0.77,1.89] |
|  | White | Ref | Ref | Ref |
| Household Size | 1-2 People | Ref | Ref | Ref |
|  | 3-5 People | 1.26 [1.05,1.52] | 1.30 [1.03,1.65] | 1.41 [1.26,1.58] |
|  | 6+ People | 1.77 [1.24,2.54] | 2.32 [1.53,3.52] | 2.17 [1.72,2.72] |
| Deprivation Index Quintile | 1 - Most Deprived | Ref | Ref | Ref |
|  | 2 | 0.74 [0.57,0.96] | 0.86 [0.62,1.19] | 1.01 [0.85,1.20] |
|  | 3 | 0.66 [0.52,0.85] | 0.72 [0.52,1.00] | 0.80 [0.67,0.95] |
|  | 4 | 0.60 [0.47,0.76] | 0.72 [0.52,0.99] | 0.80 [0.68,0.95] |
|  | 5 - Least Deprived | 0.58 [0.46,0.74] | 0.62 [0.45,0.86] | 0.70 [0.59,0.83] |

**Figure 8.**
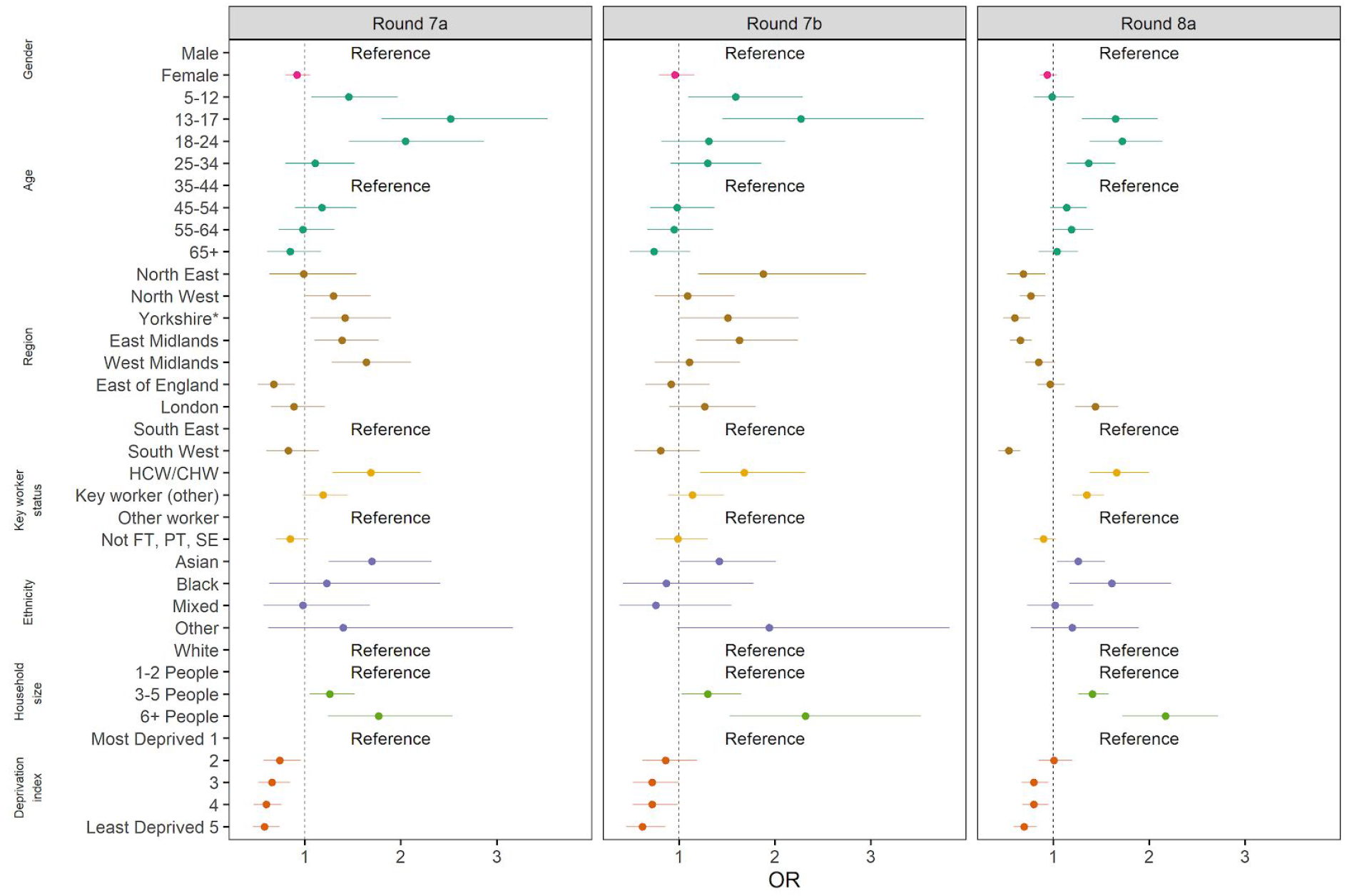
Estimated odds ratios and 95% confidence intervals for mutually-adjusted logistic regression model of swab-positivity for rounds 7a, 7b and 8a. Models were adjusted for gender, age group, region, key worker status, ethnicity, household size, and deprivation index. The deprivation index is based on the Index of Multiple Deprivation (2019) at lower super output area. Here we group scores into quintiles, where 1 = most deprived and 5 = least deprived. HCW/CHW = healthcare or care home workers; Not FT, PT, SE = Not full-time, part-time, or self-employed. *Yorkshire and The Humber.

We also found increased odds of swab-positivity among healthcare and care home workers, and other key workers, compared to other workers at 1.66 (1.38, 2.00) and 1.35 (1.20, 1.53) respectively (Table 5, Figure 8).

We observed apparent heterogeneity in prevalence across England at lower-tier local authority (LTLA) level using a nearest-neighbours statistic to generate smoothed prevalence maps (Figure 9) and maps of differences in prevalence between sequential rounds (Figure 10). In round 7b (25th November to 3rd December 2020) we found areas of higher prevalence in North East, southern parts of Yorkshire and The Humber, eastern and western parts of East Midlands, east London and surrounding areas in East of England and South East (parts of Essex and Kent). We saw a different pattern in round 8a (6th to 15th January 2021), where highest prevalence was in London and a contiguous area radiating out into East of England and South East. We also saw pockets of high prevalence in other regions. In addition, we observed high prevalence of swab-positivity across north, north-west and south-east London (Figure 11).

**Figure 9.**
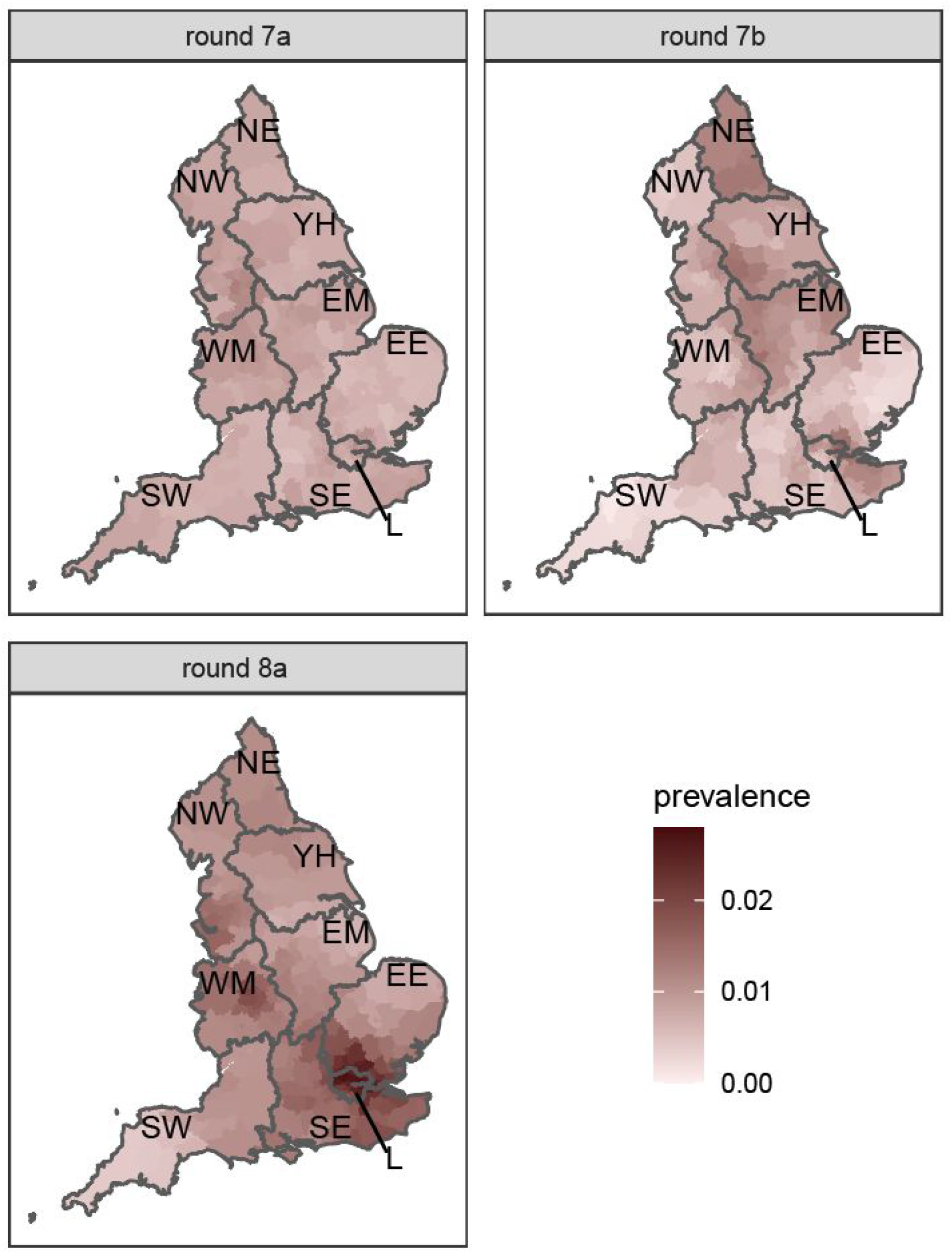
Neighbourhood prevalence of swab-positivity for rounds 7a, 7b, and 8a. Neighbourhood prevalence calculated from nearest neighbours (the median number of neighbours within 30km in the study). Average neighbourhood prevalence displayed for individual lower-tier local authorities. Regions: NE = North East, NW = North West, YH = Yorkshire and The Humber, EM = East Midlands, WM = West Midlands, EE = East of England, L = London, SE = South East, SW = South West. Data for unweighted point estimate of prevalence available in the supplementary data file.

**Figure 10.**
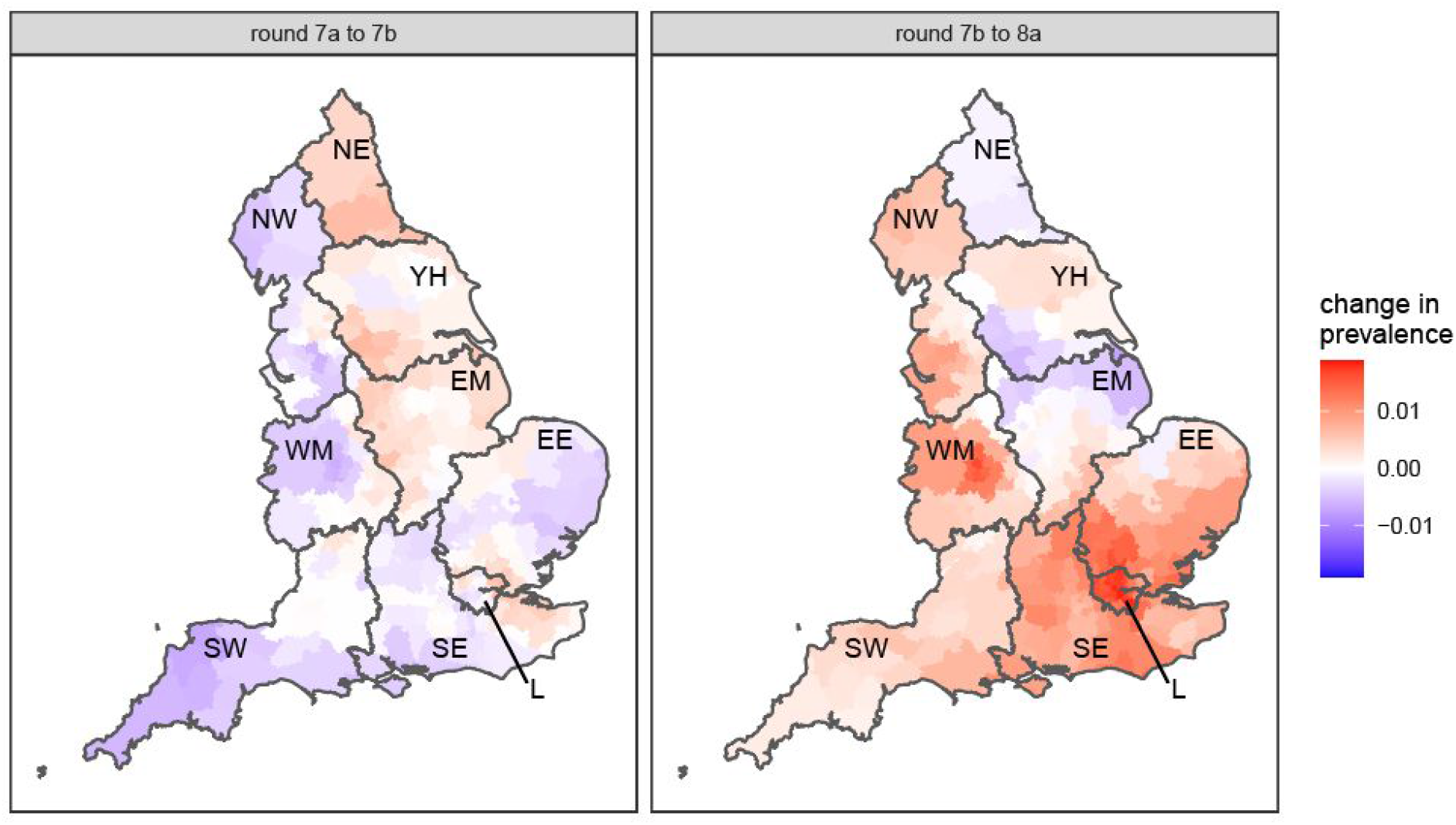
Difference in neighbourhood prevalence of swab-positivity at lower-tier local authority level from: rounds 7a, 7b, and 8a. Neighbourhood prevalence calculated from nearest neighbours (the median number of neighbours within 30 km in the study). Average neighbourhood prevalence and difference displayed for individual lower-tier local authorities. Regions: NE = North East, NW = North West, YH = Yorkshire and The Humber, EM = East Midlands, WM = West Midlands, EE = East of England, L = London, SE = South East, SW = South West.

**Figure 11.**
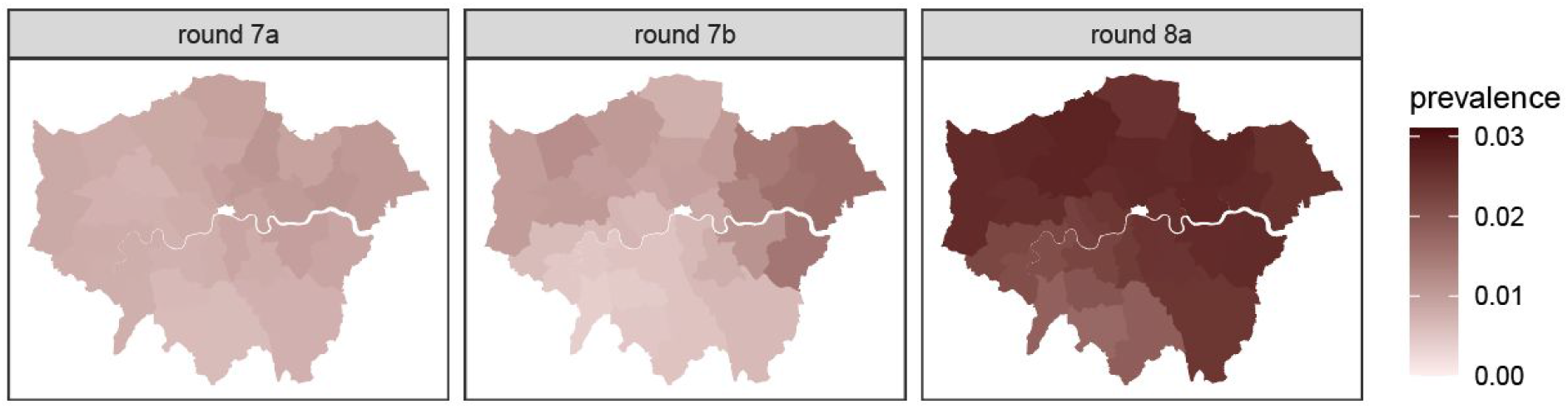
Neighbourhood prevalence of swab-positivity for rounds 7a, 7b, and 8a for London. Neighbourhood prevalence calculated from nearest neighbours (the median number of neighbours within 30 km in the study). Average neighbourhood prevalence displayed for individual lower-tier local authorities.

## Discussion

In the initial period of the third national lockdown in England, we did not observe a continued decline in the prevalence of SARS-CoV-2, as was seen in the routine surveillance data for a similar period [8]. Rather, we observed a slight initial decline followed by a plateau or possible increase, but with a weighted average prevalence substantially lower than that reported for end December and beginning of January by the Office for National Statistics Coronavirus (COVID-19) Infection Survey [5]. It is therefore possible that prevalence may have dropped substantially just prior to the start of REACT-1 round 8a.

The Facebook data presented here indicate a sharp drop in mobility in the last two weeks of 2020 followed by a return to intermediate levels during the first week of 2021. A relationship between mobility data and transmissibility was documented during spring 2020 [12,13], and we might expect the two still to be correlated: higher transmission from increased activity starting on Monday 5th January 2021 may only just be feeding into routine surveillance data. We therefore might expect to see a plateau or slight increase in routine surveillance data in subsequent days.

A key limitation in our analysis is that the trends showing a plateau or upturn in prevalence are based on data for a small number of days. However, regional prevalence trends also support there being a plateau and possible upturn (Figure 3): seven of the nine regions exhibited this pattern, while in one region (South West) there was a sharp decline. Also, the trend we report could be biased by short-term changes in testing behaviour among symptomatic individuals as they return to work. However, we obtain similar results for the constant growth model when fitting only to swab-positivity among non-symptomatic individuals (Table 2).

In the period prior to the widespread acquisition of vaccine-acquired immunity, high constant or increasing prevalence -- with high rates in at-risk age groups -- has clear downstream implications for healthcare utilization and deaths. A daily average of 950 deaths were reported in England for the seven days prior to 8th January 2021 in people who had tested positive for SARS-CoV-2 in the previous 28 days [8]. Also on 8th of January, there were 29,346 COVID-19 inpatients in English hospitals, 55% more than the maximum during the first wave [8]. If the prevalence of infections in the community does not drop substantially in the immediate future, these levels of hospitalization will lead to very high numbers of additional deaths and potentially long-term negative impact on healthcare delivery in England.

## Methods

We have published the methods of the REACT programme [14]. Briefly, between 120,000 and 180,000 people in the community in England have been taking part in the REACT-1 study at approximately monthly intervals. We invite a random sample of the population of England from the list of National Health Service patients at LTLA level (n=315). We then send a swab kit to individuals registering for the study and request they provide a self-administered throat and nose swab or parent/guardian obtain such a swab for children aged 5 to 12 years. We ask participants to refrigerate the sample for same or next day pick-up by courier for RT-PCR in a single commercial laboratory, and to complete an online or telephone administered questionnaire with information on symptoms, health and lifestyle. We estimate time trends and reproduction number (R) using exponential growth models at both national and regional levels, as well as smoothed estimates using a p-spline function [15]. We obtain estimates of prevalence of SARS-CoV-2 infection nationally and regionally, both unweighted and weighted to be representative of the population, and by age, area deprivation, occupation, household size and other characteristics. We also describe geographic variation in prevalence by LTLA using neighbourhood-based spatial smoothing as previously described [16]. We estimate odds of swab-positivity using multivariable logistic regression for a range of variables including employment, deprivation, ethnicity and household size. Statistical analyses are carried out in R [17].

We obtained research ethics approval from the South Central-Berkshire B Research Ethics Committee (IRAS ID: 283787).

## Data Availability

The datasets generated or analyzed, or both, during this study are not publicly available because of governmance restrictions.

## Data availability

Supporting data for tables and figures are available either: in this Google [spreadsheet](https://docs.google.com/spreadsheets/d/1rZD-pzU6SYEPll_bTmBzM363DJ8LYBDdV2i2QI4FZSE/edit?usp=sharing); or in the inst/extdata directory of this [GitHub R package](https://github.com/mrc-ide/reactidd).

## Acknowledgements

SR, CAD acknowledge support: MRC Centre for Global Infectious Disease Analysis, National Institute for Health Research (NIHR) Health Protection Research Unit (HPRU), Wellcome Trust (200861/Z/16/Z, 200187/Z/15/Z), and Centres for Disease Control and Prevention (US, U01CK0005-01-02). GC is supported by an NIHR Professorship. PE is Director of the MRC Centre for Environment and Health (MR/L01341X/1, MR/S019669/1). PE acknowledges support from Health Data Research UK (HDR UK); the NIHR Imperial Biomedical Research Centre; NIHR HPRUs in Chemical and Radiation Threats and Hazards, and Environmental Exposures and Health; the British Heart Foundation Centre for Research Excellence at Imperial College London (RE/18/4/34215); and the UK Dementia Research Institute at Imperial (MC_PC_17114). We thank The Huo Family Foundation for their support of our work on COVID-19.

We thank key collaborators on this work – Ipsos MORI: Kelly Beaver, Sam Clemens, Gary Welch, Nicholas Gilby, Kelly Ward and Kevin Pickering; Institute of Global Health Innovation at Imperial College: Gianluca Fontana, Dr Hutan Ashrafian, Sutha Satkunarajah, Didi Thompson and Lenny Naar; Molecular Diagnostic Unit, Imperial College London: Prof. Graham Taylor; North West London Pathology and Public Health England for help in calibration of the laboratory analyses; NHS Digital for access to the NHS register; and the Department of Health and Social Care for logistic support. SR acknowledges helpful discussion with attendees of meetings of the UK Government Office for Science (GO-Science) Scientific Pandemic Influenza – Modelling (SPI-M) committee.

